# Complex Temporal behavior Modeling for Pandemic Spread: Not a Simple Delayed Response!

**DOI:** 10.1101/2022.08.01.22278281

**Authors:** Narges M. Shahtori, S. Farokh Atashzar

## Abstract

One of the significant challenges, when a new virus circulates in a host population, is to detect the outbreak as it arises in a timely fashion and implement the appropriate preventive policies to effectively halt the spread of the disease. The conventional computational epidemic models provide a state-space representation of the dynamic changes of various sub-clusters of a society based on their exposure to the virus and are mostly developed for small-size epidemics. In this work, we reshape and reformulate the conventional computational epidemic modeling approach based on the complex temporal behavior of disease propagation in host populations, inspired by the COVID-19 pandemic. Our new proposed framework allows the construction of transmission rate (*p*) as a probabilistic function of contributing factors such as virus mutation, immunity waning, and immunity resilience. Our results unravel the interplay between transmission rate, vaccination, virus mutation, immunity loss, and their indirect impacts on the endemic states and waves of the spread. The proposed model provides a robust mathematical framework that allows policy-makers to improve preparedness for curtailing an infectious disease and unfolds the optimal time-frame for vaccination given the available resources and the probability of virus mutation for the current and unforeseen outbreaks.

## Introduction

The novel coronavirus disease, also known as COVID-19 has had a major impact on the healthcare system over the last 2 years. Series of new cases and hospitalization spikes, put intense pressure on the health care staff and resources, resulting in an estimated total loss of $323.1B and over 1 million deaths in the US alone (1).

Some of Coronavirus’s peculiar epidemiological traits and unavoidable delays in implementing the mitigation strategies, make the prevention efforts to halt the spread of the disease more challenging. In-apparent transmission through human-to-human contact when an infectious person shows no to mild symptoms and high viral loads of virus in the upper respiratory system makes Covid-19 more contagious compared to other strains such as SARS-CoV-1 and MERS-CoV (2). Particularly, epidemiological studies indicate that a significant number of carriers are asymptomatic and are unaware that they are carrying the virus (3) (4) (5).

Ongoing disease transmission and a high number of actively infected individuals result in SARS-CoV-2 virus mutation over time and new variants yet more contagious are introduced (6). These lineages contain one or more genetic mutations that differentiate them from the virus variant that is already circulating in the host population and may cause the vaccination to become less effective. For instance, in November 2021 studies shows *B*.1.1.529 variant also known as omicron, which later resulted in unprecedented waves in many countries, has the ability to escape antibody immunity induced by the existing vaccines (7) (8) (9). In addition, imperfect implementation of control strategies and failure to diagnose the symptoms of the disease or its new variants, specifically at the early stage results in multiple surges in new cases (10), as has been observed in the last two years. This would call for more holistic modeling that matches the complex behavior of prolonged pandemic crises in a connected society, going beyond small-scale epidemic modeling.

The rapid advances in vaccine technology and the progress of medical sciences have dramatically enhanced our capabilities in combating viruses (11) (12). A recent study concluded that since 1924, vaccination has eliminated a large part of the infectious diseases and vaccines have successfully prevented more than 170 million cases of diphtheria, measles, and childhood diseases in the United States (13). Yet, studies warn us about the emergence of new human infectious diseases and outbreaks in the near future. Particularly, in the wake of climate change, empirical evidence anticipate perturbation in the structure of ecosystems, food supplies, and the distribution of biodiversity. Studies demonstrate that the major ecosystem shifts influence the dynamics of infectious disease spreading and the omnipresence of outbreaks (14) (15) (16). In fact, new pandemics may happen in the near future and COVID-19 might be one of the new infectious diseases that we experience in the upcoming years.

The major challenge when a virus circulates in a host population is to detect the outbreaks as they arise in a timely fashion and implement the appropriate preventive policies. A critical question, however, is what type of management policies can be applied to effectively control the spread of infectious disease and reduce the financial burden of emerging infectious diseases on the healthcare system and subsequently reduce the mortality rate. For instance, despite the worldwide efforts over the last two years to curtail Covid-19, it is still considered a public health emergency. The ongoing transmission of the virus across the globe results in virus mutation and the creation of more contagious variants. These new virus variants make it more challenging to curtail the spread of the disease.

Contact tracing and isolation are two main strategies, proper implementation of which can slow down the chain of virus transmission when vaccination is not immediately accessible to the mass population. However, implementation of those preventive strategies with no delays is rarely achievable and it heavily depends on the socioeconomic status of the host population and can be a major burden for societies with limited healthcare access (10) (17). During the coronavirus pandemic, public and private health authorities utilize different mitigation strategies such as contact tracing, isolation, and mass COVID-19 testing to curtail the outbreak.

In practice, implementation of control measures in a highly connected society with no delays, detecting the most optimal strategies, large-scale optimal resource allocation, and enforcing preventive protocols within an effective timeframe when patients outnumber the health care staff and the first responders, have not been feasible in most regions worldwide. For instance, nine months after the World Health Organization (WHO) announced COVID-19 as a pandemic, only a few countries were utilizing contact tracing effectively (18). It is imperative to notice that studies of other recent outbreaks such as Ebola epidemics in West Africa also indicate that the spread of disease was effectively controlled once preventive protocols were improved and adequate resources were allocated to reduce the time delay in identifying and tracing newly infected individuals (19) (20).

In this regard, computational and probabilistic models that formulate the statistics of virus spread propagating among various clusters of a networked society are playing an invaluable role in providing insight into the stated problems and helping decision-makers, governments, and stakeholders to implement appropriate strategies. A better understanding of the impact of delays and efficacy of the conducted mitigation at different stages of disease propagation, in addition to better prediction of the effects of potential future mutations and the changes in the status of immunity resilience, are unmet needs for the control of a pandemic-level spread and can be crucial to avoid the disabling socioeconomic pressure on many societies, caused by the virus, for example, in future unforeseen outbreaks and pandemics.

Motivated by the above-mentioned facts, in the literature, there has been a surge of efforts in the development of various computational models. Such models provide a state-space representation of the dynamic changes of various sub-clusters of a society based on their exposure to the virus and are mostly developed for small-size epidemics. For example, the commonly used method *susceptible − infected − recovered*, named *SIR*, models the connection and disease transmission between susceptible, infected, and recovered groups in a host population. More advanced models such as *susceptible − exposed − infected − recovered* (*SEIR*), include an intermediate dynamical state for the exposed group to better model the interaction between sub-population during the course of an epidemic. Specifically, individuals in the exposed state (*E*) incubate the virus for a certain period of time before becomes infectious. They are considered non-symptomatic and non-infectious during this period. In some literature, additional states are incorporated to the classical *SEIR* and *SIR* models to further enhance the modeling of the complex nature of disease spreading (2) (21) (22) (23) (24).

Going beyond the above-mentioned classical models, researchers tried to incorporate the mitigation strategies, immunity loss, and demographic effects into the mathematical infectious disease modeling and assess the effects on the disease transmission rate. In this regard, Radulescu *et al*. have enhanced the SEIR model by assembling an age-compartmental design and incorporating social mobility dynamics, to numerically study the disease progression in a small college community scenario when social mobility restrictions are enforced (25). In another effort, Bjørnstad *et al*. incorporate demographics and immunity loss into the classical *SEIR* model to assess endemic states in the presence of continuous recruitment into susceptible populations (26). However, the physics of transmission rate and average contact rate with respect to the model dynamics are disregarded in the proposed models. In a homogeneous population, we define the transmission rate (*β*), as *β* = *pω* (27) (15). *p* is the probability of disease transmission and an individual makes contact with the infected population (*I*) with the rate of *ω*. In reality, social mobility restrictions, mortality, and demographics features directly impact the average contact rate (*ω*), *p*, and subsequently *β* and thus the number of new cases. The models designed in the literature often oversimplify the interdependent effects among *ω, p*, and model states (see in (26) and (25)). Thus, such models often fail to take into account adequate factors that contribute to the magnitude of the epidemics and as a result, they cannot provide the needed accuracy in the estimation of disease spread, especially at the large scale such as a pandemic.

In addition to the above, there are limited works to realistically incorporate the delays corresponding to the implementation of mitigation strategies and lack of identifiability, as part of the state-space infectious disease modeling (28) (29) (10) (30). In an effort to assess the consequences of delays and incomplete identification of infectious individuals, Young *et al*. proposed a mathematical framework that considers the average transition time from one state to another as a form of a constant delay (10). However, the proposed model fails to capture the probabilistic nature of transition when an exposed individual incubates the virus for *σ* unit-time. For example, the work presented in (10) assumes that all of the susceptible individuals that contact with infectious individuals at time *t − σ* acquire the disease at time *t*, and therefore, the probabilistic effect of intermediate dynamical state *E* (which directly impacts the spread of disease) is not observed in the model.

in this paper, to bridge the gap between the observed reality of large-scale and long-term disease progression in a host population and currently utilized infectious disease frameworks, we redefine the computational representation of transmission rate when the exposed individuals incubate the disease for a period of time before becoming infectious. Particularly, we formulate a new dynamical interaction behavior among *S, E*, and *I* states, inspired by the observations made during the course of COVID-19 pandemic. We introduced a novel state-space model called *Susceptible*–*exposed*–*infected*–*quarantine*–*recovered − dead* (*SEIQRD*), which takes into account both temporal event-based, and probabilistic features of transitions across various states. Specifically, we model the physics of transition across the various state using the homogeneous Poisson point process concept, using which arrival and departure into/from a state are considered random events that occur during a given time interval. In this work, we shed light on the critical questions pertaining evolution of transmission rate when control measures, such as mass vaccination are implemented. Our simulations unravel the interplay between transmission rate, vaccination, virus mutation, and their indirect impacts on the endemic states and waves of the spread. Our unique mathematical framework allows us to objectively evaluate and identify the optimal management policies required to effectively curtail the spread of infectious diseases. Furthermore, our novel model provides a robust mathematical framework that allows policy-makers to improve preparedness for curtailing an infectious disease and unfolds the optimal time-frame for vaccination given the available resources and the probability of virus mutation for the current and unforeseen outbreaks. Also, it enables the policymaker and health organizations to predict and extrapolate the outcome of the different mitigation strategies. Using the proposed method, health authorities will have a powerful and flexible framework to objectively conceptualize and predict the endemic state in the context of mitigation intervention and deployment of vaccination.

### Background on Modeling and Outstanding Research Questions

Over the years, various methods have been developed to address fundamental questions pertaining to the evolution of the infectious disease in a host population and the associated risks related to reactive and proactive management policies. Computational infectious disease models allow detecting the surge of new cases and the emergence of outbreaks at an early stage. To this end, several mathematical approaches have been introduced in the literature which can be categorized into two general classes (11): statistical approaches and state-space methods.

### Conventional Statistical Models At A Glance

Statistical approaches, such as regression models and Bayesian methods are utilized to predict the emergence of outbreaks by elucidating the hidden patterns and extrapolating the observed historical data. In this regard, regression-based models have been commonly used for example to predict the emergence of influenza outbreaks at early stages. One of the known models developed to estimate the average influenza mortality using the regression method is proposed by *Serfling* (31). The model proposed by *Serfling*, given in (1), incorporates the seasonal behavior, historical data on influenza, and reported cases in order to predict the emergence of new outbreaks (32).

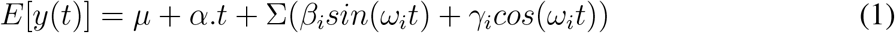

In (1), *µ* represents the expected value of the reported cases based on the historical data and ∑(*β*_*i*_*sin*(*ω*_*i*_*t*) + *γ*_*i*_*cos*(*ω*_*i*_*t*)) accounts for cyclical trends of the influenza virus. The simplified version of *Serfling*’s approach is currently utilized by the Centers of Disease Control (CDC) in the US, Australia, France, and Italy for the identification of influenza outbreaks (11). Over the years, there have been major efforts to enhance *Serfling*’s model by incorporating the noise into the predictions and accounting for uncertainties (33). For example, researchers conduct several approaches such as generalized additive models (GAMs) (34) and wavelet transform (35) to address discontinuity in the historical data for computing the mean and standard deviations in 1. In another line of research related to statistical methods, researchers implement the hidden Markov model (HMM) (36) and Markov Chain Monte Carlo (MCMC) (37) to incorporate the hidden patterns (states) of the disease spread and forecast the outbreak. In this method, the observed cases, *y*(*t*), is associated with a set of hidden variables, *x*(*t*), which it determines the conditional probability of *y*(*t*)|*x*(*t*) *∼ f*_*i*_(*y*(*t*); *θ*_*i*_). It should be noted that some of the proposed statistical models for influenza are utilized to predict the emergence of other infectious disease outbreaks (38) (39) (40) (41).

### State-Space Model

State-space models are used to predict the evolution of outbreaks over time and assess the effectiveness of mitigation strategies. The conventional state-space model that allows for the incorporation of the relevant contributing factors of infectious disease spread was proposed by *Kermack and McKendrick* (42). The state-space *SIR* (*susceptible − infected − recovered*) model proposed by *Kermack and McKendrick* has been widely used to predict new outbreaks and model infectious disease spread (43). In this context, the host population is divided into different groups based on the state of their health and their interactional status with the infected sub-population. (2) represents, *SIR* state-space model proposed by *Kermack and McKendrick* (42).

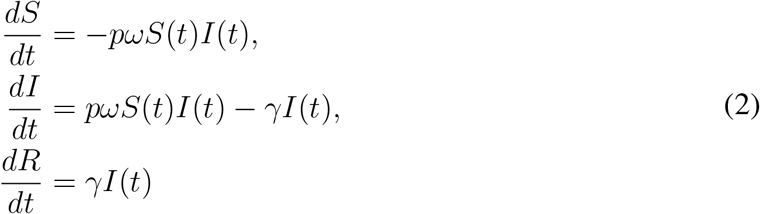

In the *SIR* model, given above, at time *t*, infected sub-population (*I*(*t*)) make contacts with susceptible sub-population (*S*(*t*)) with rate *ω* and a susceptible individual contract the disease with probability *p*. Thus, an infected person transmits the disease to *pωS* susceptible individuals at a unit of time. *−pωSI* term indicates the number of susceptible individuals who enter the infectious group *I*. Then, infected individuals move to the recovered/dead sub-population (*R*) within *γ*^*−*1^ unit time (44). In this framework, the *R* compartment is considered the sub-population that cannot get reinfected. In addition, the size of the host population (*N*) is assumed to remain constant throughout the outbreak and the host population is considered to be homogeneous, i.e. individuals in the host populations have an equal probability to make contact with others and every susceptible individual has the same probability of becoming infected.

Over the last few decades, the *SIR* model is enhanced by adding another state named exposed, *E*. The model is also known as *susceptible − exposed − infected − recovered* or in short *SEIR*. This model is widely used in the literature when exposed individuals incubate the virus for *σ*^*−*1^ unit time. (3) represents the state-space model of *SEIR* and formulates transitions across various states mathematically.

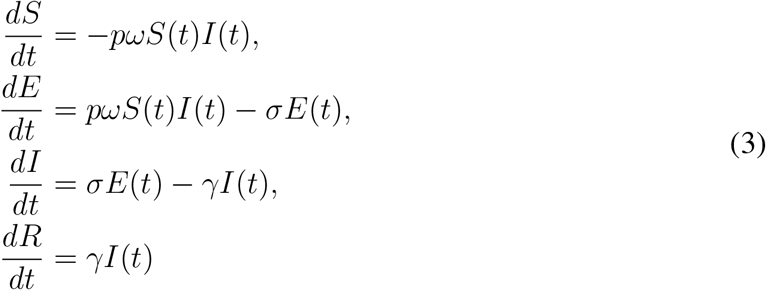

In the *SEIR* model, presented above, it is assumed that susceptible individuals leave the *S* group when they become in contact with infectious individuals (*I*), and contract the disease with a transmission probability of *p*. The exposed individuals (*E*) are considered non-symptomatic and non-infectious during the incubation period. Exposed individuals incubate the disease for *σ*^*−*1^ unit time before moving to the infectious state at rate *σ*. Then, infected individuals enter the recovered/dead state (*R*) after *γ*^*−*1^ unit time.

The classic *SEIR* model is suitable to simulate and predict small-size outbreaks. However, there are 3 major issues with the conventional *SEIR* models concerning outlining a realistic realization of large-scale epidemics or pandemics.

- **Problem (1)** In the classic *SEIR* model, it is assumed that individuals who contact an infectious person contract the disease with the probability of *p* and leave the *S* state with the rate of *pω*. However, this assumption does not take into account that a portion of the exposed population would not contract the disease and they return to the *S* population. In reality, the interactional status between the *E* and the *S* states is directly controlled by the health authorities and policymakers for curtailing the disease spread through tracing the detected exposed individuals (also known as “contact tracing”). Recruitment of exposed individuals who do not contract the disease to susceptible sub-population after the incubation period is an important dynamic pattern that plays an integral part in the spread of disease and has been disregarded in the literature.
- **Problem (2)** Over the last three years, it has been shown that implementation of preventive protocols and vaccination are associated with delays. The delays pertaining to mitigation strategies are another important dynamic that directly impacts the behavior of the system at endemic state. There has been limited work to incorporate the delays corresponding to the implementation of mitigation strategies, as part of the state-space infectious disease modeling, e.g., (28) (29) (10) (45) (46). However, the proposed models oversimplify the probabilistic nature of transition and the temporal inter-dependency between *p, ω, S*, and *I* when an exposed individual incubates the virus for *σ* unit-time.
- **Problem (3)** Virus mutation and vaccination directly impact the probability of transmission, *p*, and subsequently the number of new cases in the host populations. However, mutation and development of immunity resilience against the virus are overlooked as contributing factors in the literature when modeling the disease spread.

## Results and Discussion

In this paper, we propose a new computational model, going beyond classic *SEIR* modeling, using a homogeneous Poisson point process for the first time that addresses the previously mentioned issues. The proposed model in this paper takes into account,(a) the interactional status between the *E* and the *S* states when exposed individuals do not contract the disease, (b) the inter-dependency between *p, ω, S* and *I* states and (c) the effects of mutation and development of immunity resilience in the society. Such a model can be imperative when controlling a long pandemic in a mega population that echoes waves of mutation and spread.

### Reformulating *SEIR* model

To address critical issues with the conventional *SEIR* model such as oversimplification of the interplay between exposed, susceptible and infectious states, and the inter-dependency between *p, ω, S*, and *I*, we propose a novel framework for the *SEIR* model which utilizes the Poisson point process to define transition across various states. We consider the transitions between *E → I* and *I → R* as an arrival Poisson point process, meaning an individual in the host population arrives at a new state (given the health status) within a predefined period. We can utilize the Poisson point process concept to model this behavior because the average transition period is known but the exact arrival time to a new state is random. Therefore, we can formulate the transition rate as the average number of arrivals (events) given the transition period using (13).

We assume a susceptible individual who becomes in contact with an infectious person leaves the susceptible group (*S*) with rate *ω*. The newly exposed individuals (*E*), *ωS*(*t*)*I*(*t*), at time *t*, are considered non-symptomatic and non-infectious. Notably, only a fraction of exposed individuals become infectious. The exposed individuals who contract the disease with probability *p* move to the infectious state *I* with rate *λ*_*EI*_ after incubating the disease for *σ* unit time. *λ*_*EI*_ represents the average number of arrivals to state *I* given the transition period of *σ*. The exposed individuals who do not contract the disease after *σ* unit time, return to the susceptible population with rate *λ*_*ES*_. Specifically, *λ*_*ES*_ represents the average number of individuals who return to state *S* given the transition period of *σ*. Then, infected individuals enter the recovered/dead state (*R*) with rate *λ*_*IR*_ after *γ* unit time. Similar to the previously introduced rates, *λ*_*IR*_ represents the average number of individuals moving to *R* state given *γ* unit time. (4) represents the computational framework for the proposed model.

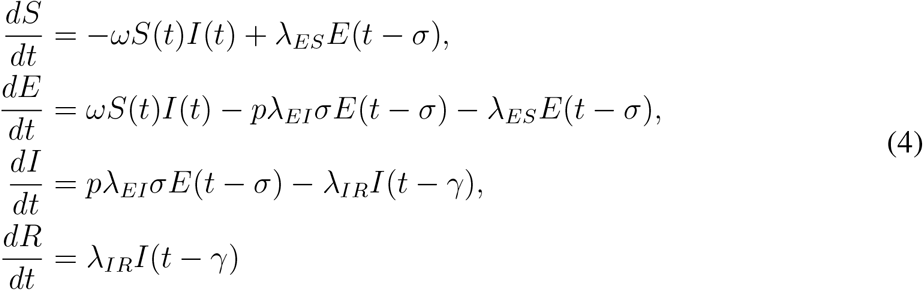

This framework formulates the dynamic changes of various sub-clusters of the host population based on their exposure to the virus when the exposed group is an intermediate step between the susceptible and the infectious states. The proposed model reconstructs the *S → E → I* transition, by considering the fact that changes in the susceptible population occur only when individuals get exposed to the virus and not when they contract the disease. Furthermore, the proposed model allows for construction *p* as a function of contributing factors. We propose a unique model for *p* by taking into account the relevant factors such as virus mutation, vaccination, and immunity loss in section.. Such a model can be utilized to simulate the future waves of pandemics depending on an assumed temporal expectation of the mutation. Also, the new formulation allows for the evaluation of the disease spread in various societies and sub-societies with different immunity responses and vaccination profiles.

### Probabilistic temporal event-based disease progression model

To address the critical questions mentioned in section and assess the impact of vaccination objectively, we proposed a novel mathematical framework that (a) takes into account the interdependent relations between transmission rate (*p*), contact rate (*ω*) and immunity loss (*λ*_*R*_*SR*(*t − α*)), and (b) formulates the dynamical temporal event-base interactional status across various states. The proposed model, *Susceptible-exposed-infected–quarantine–recovered-dead* (*SEIRQD*) divides the host population into six groups, depending on the state of an individual’s health and whether or not they are exposed to the virus through an infected person. Figure.1 depicts the summary of state transitions in the *SEIQRD* model.

**Figure 1:**
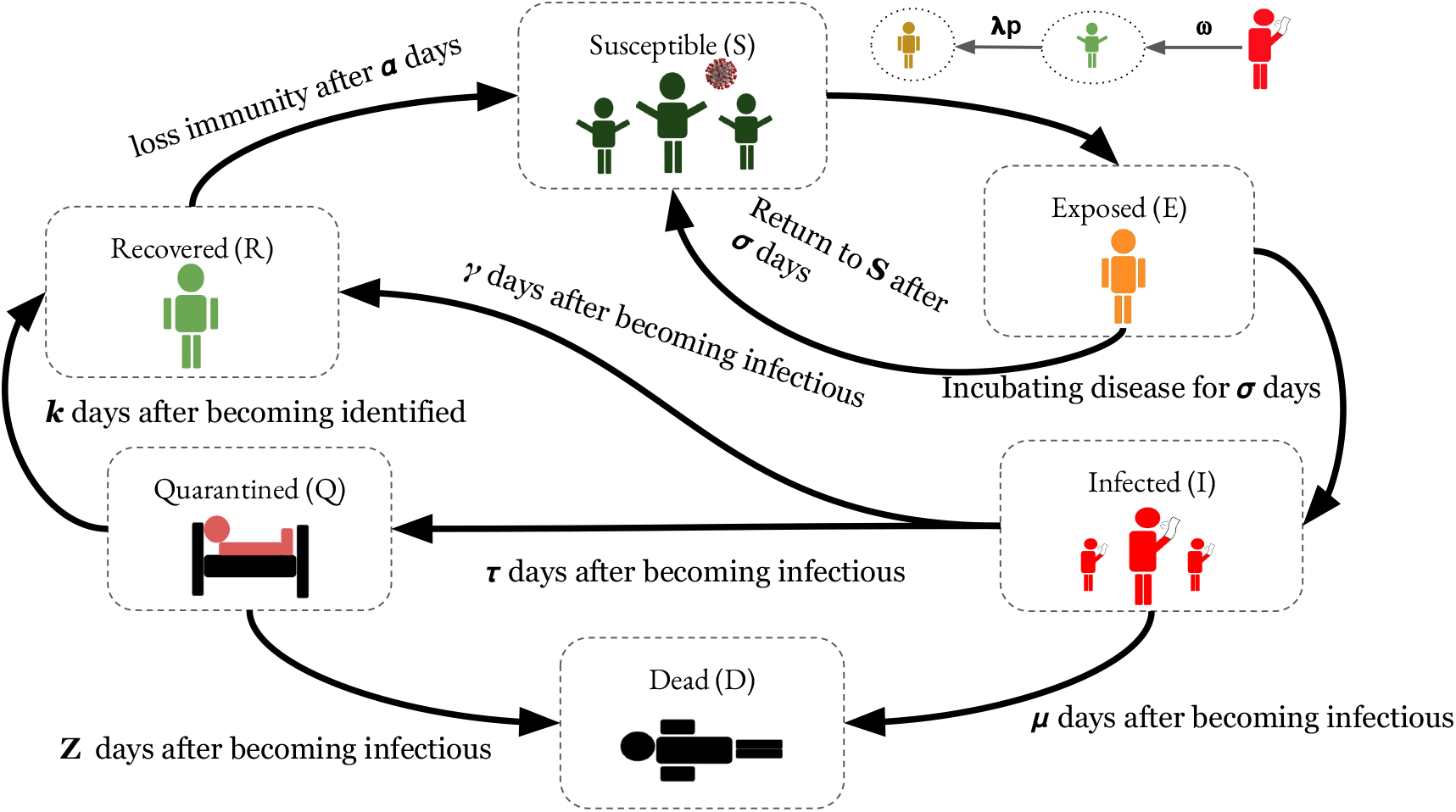
A simplified summary of the proposed model (*SEIQRD*) and the corresponding spread chain: Infectious individuals become in contact with healthy individuals (*S*) with rate *ω*. A fraction of exposed ones acquire the disease after incubating the disease for σ unit time and move to infectious state (*I*) with rate *pλ*_*EI*_. Other exposed individuals that do not contract the disease return to the susceptible population with rate *λ*_*ES*_. Once an exposed individual becomes infectious, they would have 3 possibilities: 1. they are identified and placed into isolation, state *Q*, 2. they recover and enter state *R*, or 3. they pass away (state *D*) and get removed from the spreading cycle. Similarly, infected individuals who are identified, either recover or pass away. Finally, recovered individuals are recruited to the susceptible state due to waning immunity after *α* unit time with rate *λ*_*RS*_.

Infectious individuals (*I*) become in contact with healthy individuals (*S*) at rate *ω*. A fraction of exposed ones (*E*) acquires the disease after incubating the disease for *σ* unit of time and move to an infectious state (*I*) with rate *pλ*_*EI*_. In this paradigm, the term *ωS*(*t*)*I*(*t*) represents the number of individuals who become in contact with infected individuals at time *t*. A fraction of those who are exposed to the virus at time *t − σ* (i.e. *E*(*t − σ*)) contract the disease after incubating the virus for a period of *σ* unit time. During this period they are considered non-symptomatic and non-infectious. Once an exposed individual becomes infectious, they would have 3 possibilities: (1) they are identified and placed into isolation, state *Q*, (2) they recover and enter state *R*, or (3) they pass away (state *D*) and get removed from the spreading cycle. The exposed individuals who do not contract the disease after *σ* unit time, return to the susceptible population with rate *λ*_*ES*_. Infected individuals who are identified, either recover or pass away. Finally, recovered individuals are recruited to the susceptible state after *α* unit time with rate *λ*_*RS*_.

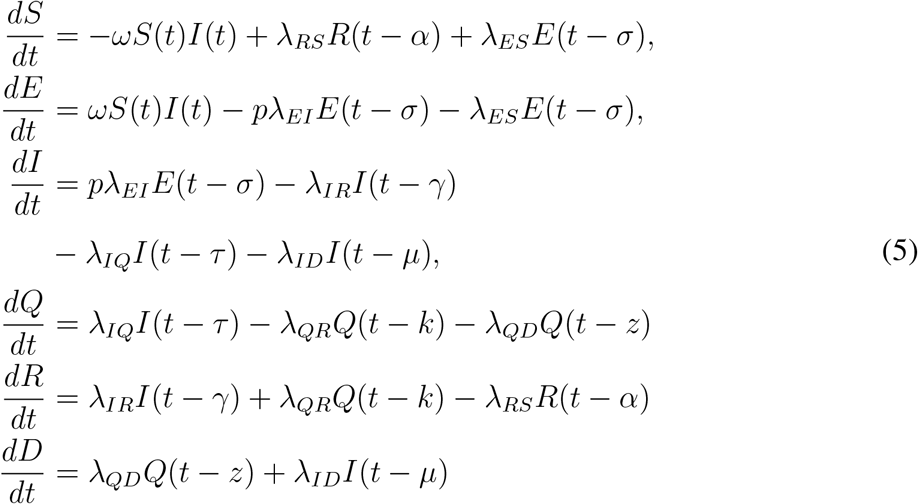

(5) represents the proposed *SEIQRD* mathematical model and Table.1 delineates the model’s parameters. Individuals move to a new group (e.g. arrival to a new group) after staying in the current group for *y* unit time. We formulate this behavior using the homogeneous Poisson process because the average transition time is known but the exact arrival time to the new state is random. Per (12) and (13), the average inter-arrival duration between *i −* 1^*th*^ and *i*^*th*^ moments in a Poisson point process with rate *λ* forms an exponential distribution, with the expected value of *λ*^*−*1^. Therefore, we define the *E → I, E → S, I → D, I → Q, I → R, Q → R, Q → D* transitions as follows:

- *E → I*: During *σ* unit time, on average 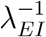 exposed individuals who contracted the virus with probability *p* undergo a transition to state *E*. This implies that exposed individuals who become infectious enter state *I* with *σ* unit time delay with the average rate of *λ*_*EI*_ which is reflected as *pλ*_*EI*_*E*(*t − σ*) in (5).
- *E → S*: During *σ* unit time, on average 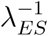 exposed individuals that do not contract the virus undergo a transition to state *S*. This implies that exposed individuals who are not infectious return to state *S* with *σ* unit time delay with the average rate of *λ*_*ES*_ which is reflected as *λ*_*ES*_*E*(*t − σ*) in (5).
- *I → D*: The infected individuals at time *t − µ* pass away with the average rate of *λ*_*ID*_ after remaining contagious for *µ* unit time. Particularly, the arrival at state *D* between the (*i −* 1)^*th*^ and (*i*)^*th*^ moments is an exponential random variable with rate 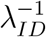. This transition is indicated in (5) as *λ*_*ID*_*I*(*t − µ*).
- *I → Q*: The average number of infectious who are identified during *τ* unit time is 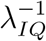. Therefore, the infectious individuals at time *t − τ* undergo a transition to state *Q* with the average rate of *λ*_*IQ*_ at time *t*. This behavior is modeled as *λ*_*IQ*_*I*(*t − τ*) in (5).
- *I → R*: During *γ* unit time, the average number of unidentified infected individuals who are recovered is 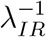. As a result, infectious individuals at time *t − γ* move to state *R* with the average rate of *λ*_*IR*_ at time *t*. This behavior is modeled as *λ*_*IR*_*I*(*t − γ*) in (5).
- *Q → R*: Term *λ*_*QR*_*Q*(*t − k*) in (5) indicates the number of identified infected individuals at time *t − k* who are recovered with the average rate of *λ*_*QR*_ at time *t*.
- *Q → D*: Term *λ*_*QD*_*Q*(*t − z*) defines the changes in quarantine population at time *t*. These individuals enter group *Q* at time *t − z* and pass away at the average rate of *λ*_*QD*_.
- *R → S*: Individuals who are recovered at time *t − α* maintain the immunity against the disease for *α* unit time and immunity wanes with the average rate of *λ*_*RS*_.

**Table 1:** Parameters of SEIQRD model

| Description | Parameter |
| --- | --- |
| Average contact rate per unit time | $\omega$ |
| Disease transmission rate | $p$ |
| Lucky rate | $\lambda_{ES}$ |
| Loss of immunity rate | $\lambda_{RS}$ |
| Exposure rate | $\lambda_{EI}$ |
| Identification rate | $\lambda_{IQ}$ |
| Mortality rate of the unidentified infectious | $\lambda_{ID}$ |
| Recovery rate of the unidentified infectious | $\lambda_{IR}$ |
| Mortality rate of the identified infectious | $\lambda_{QD}$ |
| Recovery rate of the identified infectious | $\lambda_{QR}$ |
| Incubation period | $\sigma$ |
| Time elapsed between recovery and loss of immunity | $\alpha$ |
| Time elapsed between infection and identification | $\tau$ |
| Time elapsed between identification and recovery | $k$ |
| Time elapsed between identification and death | $z$ |
| Time elapsed between infection and death without being identified | $\mu$ |
| Time elapsed between infection and recovery without being identified | $\gamma$ |

The proposed model in this section maps out the interplay between various states by taking into account the complex temporal interaction and inherent dynamics. However, to assess the impact of vaccination on the transmission rate and ultimately the disease propagation, we need to define *p* as a function of relevant factors i.e. virus mutation, vaccination, and immunity loss. In the next section, we propose a framework that can be integrated with the proposed model to represent the response of the system to mutation, vaccination, and immunity loss. Such a model can be utilized to simulate the future waves of pandemics depending on an assumed temporal expectation of the mutation. Also, this allows for the evaluation of the disease spread in various societies and sub-societies with different immunity responses and vaccination profiles.

### Disease transmission rate

We formulate *p*(*t*) by incorporating virus mutation, waning immunity, and population’s immunity resilience against virus as given below:

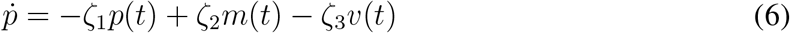

(6) represents the mathematical expression of *p*(*t*). In this model, *v*(*t*) reflects the host population’s immunity resilience against virus boosted by vaccination efforts, and *m*(*t*) indicates the communication of virus variants from one person to another. *ζ*_1_ represents the overall rate at which natural immunity is produced against the virus in the host population. *ζ*_2_ is the rate at which the virus spreads given the circulating variant at time *t*, and *ζ*_3_ represents how well the vaccination efforts are implemented. We use (7) to generate a sigmoid shape curve for *v*(*t*). Particularly, this function generates an *S-shape* growth curve in which immunity resilience boosted by vaccination increases slowly initially, and when mass vaccination becomes available approaches an exponential growth rate.

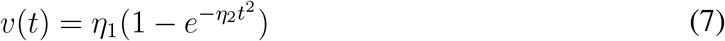

Virus variant’s contagion rate is modeled using (8). We utilize multi-step functions as indicated in (8) to take into account virus mutation occurrence and model the corresponding contagion rate.

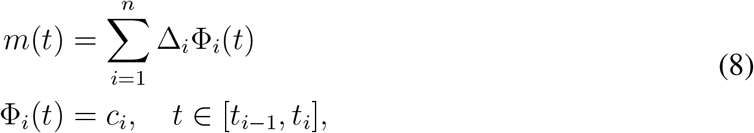

Infected individuals who are recovered develop natural immunity against the virus which results in a reduction of *p*(*t*) over time. We formulate this behavior by adding *−ζ*_1_*p*(*t*) term in (6).

### Interplay Between Vaccination, Transmission Rate, and Infectious Group

Here, we conduct a systematic simulation study to evaluate the behavior of the proposed model constructed in (5) to generate a realistic realization of infectious disease spread and evaluate the impact of mass vaccination in combating the disease propagation. In our simulation we assume, population size *N* is 8*e*6, number of initially infected individuals equals to *I*_0_ = 1, and the initial transmission rate at time *t* = 0 is *p*(0) = 3.2*e−*1. We considered average contact rate, *ω ∈* [0.5, 0.7]. To account for the communicable disease, we considered a relatively high exposure rate, *λ*_*EI*_ = 2.8*e−*2, and a short incubation period, *σ* = 5 unit time. Particularly, *λ*_*EI*_ = 2.8*e−*2 indicates that on average 35 individuals who contracted the disease with rate *p* move to infectious state within *σ* unit time. Exposed individuals who did not contract the disease after *σ* = 5 unit time were recruited to the susceptible population with rate *λ*_*ES*_ = 3.4*e−*2. Then, infected individuals either got recovered with an average rate *λ*_*IR*_ = 1.4*e−*2 within *γ* = 14 unit time or passed away with an average rate *λ*_*ID*_ = 8*e−*4 within *µ* = 15 unit time. Other infected individuals were identified with an average rate *λ*_*IQ*_ = 1.12*e−*2 within *τ* = 5 unit time. The identified infectious individuals were placed into isolation and therefore could not contact a susceptible individual. The quarantined infectious individuals stayed in the isolation for *k* = 21 unit time until they fully recovered and entered *R* state with an average rate *λ*_*QR*_ = 1.8*e−*3. The rest of the quarantined population passed away after *z* = 25 unit time at average rate *λ*_*Q*_*D* = 6*e−*4. We assumed natural immunity waned after *α* = 200 unit time and recovered individuals were recruited to the susceptible population at average rate *λ*_*RS*_ = 7.2*e−*3.

We utilized (6) to enforce the time-variant transmission rate. We considered three virus mutations in the first 500 unit time. Specifically, new virus variants were introduced at time *t* = [145, 260, 360]. Figure.2b shows the virus mutations and the corresponding contagion rate. We formulated the contagion rate using (8) as follows:

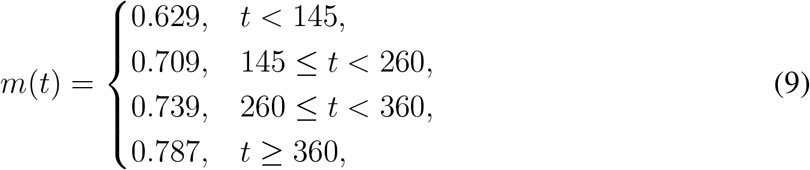

**Figure 2:**
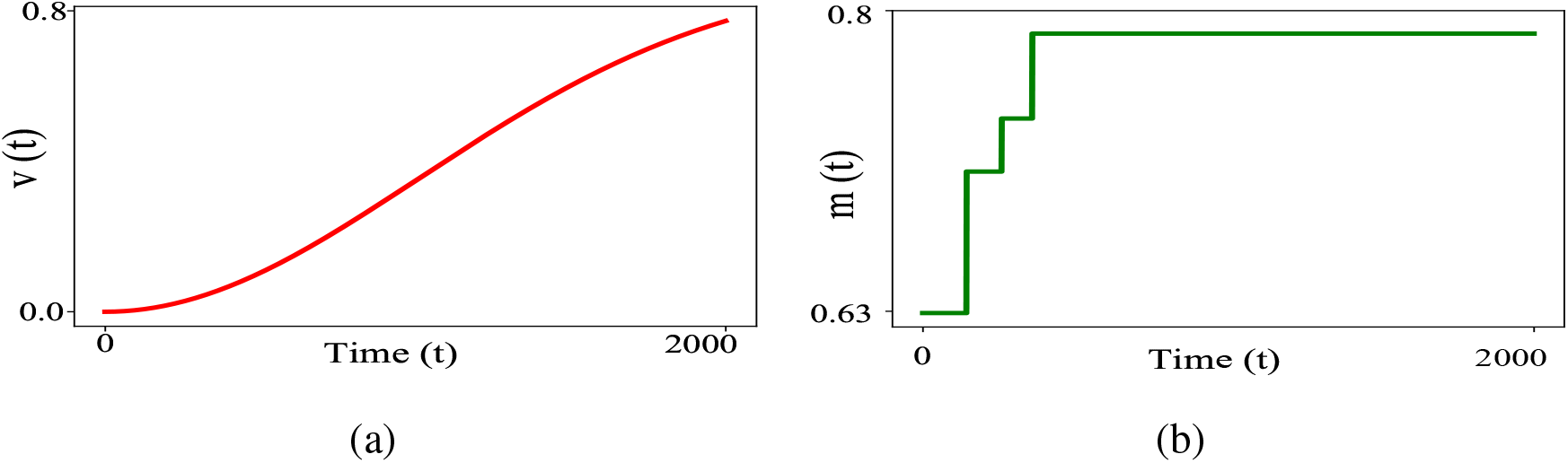
(a) Simulated realization for vaccination function (*v*(*t*)) which represents the host population’s immunity against virus. (b) Simulated realization of mutation function (*m*(*t*)). (7) is utilized with parameters *η*_1_ = 9*e* − 1 and *η*_2_ = 1*e* − 4 to generate *v*(*t*) function. (8) is utilized to generate a realization of virus mutation (*m*(*t*)) due to ongoing disease transmission results.

To objectively assess the impact of vaccination on *p*(*t*) and subsequently the control of disease spread, we considered *ζ*_3_ as the control measure. Particularly, per (6), three factors directly impact the transmission rate (*p*(*t*)), *ζ*_1_, *ζ*_2_, and *ζ*_3_. *ζ*_3_ is the external factor that can be utilized by policy makers to control the transmission rate *p*(*t*) and subsequently the progression of the disease. Figure.2a illustrates the percentage of the host population that is vaccinated over the course of 2000 unit time span. We tuned *η*_1_ = 9*e−*1 and *η*_2_ = 1*e−*4 to generate a scenario where available resources allow the policy makers to vaccinate 80% of population over 2000 unit time span. *ζ*_1_ and *ζ*_2_ are determined by the nature of virus and we assumed *ζ*_1_ = 6.8*e−*1 and *ζ*_2_ = 9.2*e−*1. We performed *9* different scenarios to assess the effectiveness of vaccination.

Figure.3a and 3b illustrate the evolution of *p* when *ζ*_3_ *∈ {*0.4 + *x ≤* 0.8, *x* = .05*}* and *ω ∈* [0.5, 0.7]. During the first 500 unit time, *p* reaches its maximum value 1 due to multiple virus mutations (Figure.2b) and insufficient immunity resilience against virus (term *ζ*_3_*v*(*t*) and *ζ*_1_*p*(*t*) in (6)). However, *p* reduces when host population’s immunity resilience increases and eventually converges. As shown in Figure.4a, the biggest jump in the number of new cases occurs when *p*(*t*) reaches its maximum value 1 for both *ω* = 0.5 and *ω* = 0.7, which indicates the direct relationship between the number of cases and *p*. Figure.5c and 6c indicate the relation between *ζ*_3_ and size of infectious group (*I*). It indicates that higher value of *ζ*_3_ results in fewer infectious cases. Therefore, size of infectious group (*I*) has inverse relation with *ζ*_3_ and direct relation with *p*. Our simulation indicates that the disease spread is halted entirely when *p*(*t*) converges to 0(or *{ζ*_3_ = 8*e*^*−*1^, *ω* = 0.7*}* and *{ζ*_3_ *∈* [0.75, 0.8], *ω* = 0.5*}*). For cases that *p*(*t*) *>>* 0 (Figure.3a and 3b), disease continues to circulate in the host population and the magnitude of *p*(*t*) and *ω* directly impact the size of infectious group (Figure.4a).

**Figure 3:**
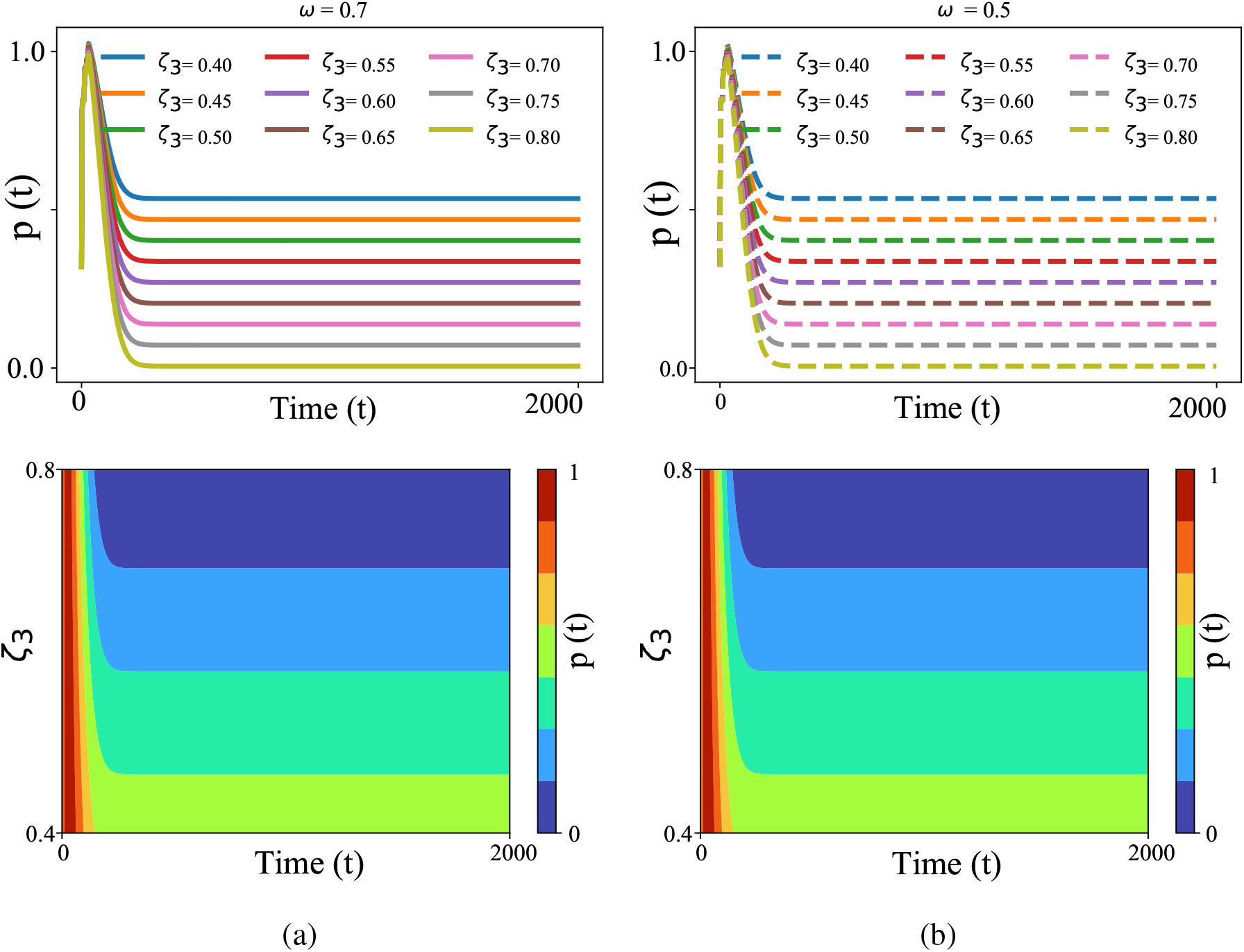
Evolution of transmission rate, *p*(*t*), over time with respect to performance of vaccination implementation (*ζ*_3_) when average contact rate, *ω*, is 0.7 (Figure.3a) and when *ω* = 0.5 (Figure.3b). *ζ*_3_ plays an input control role which defines how effective mass vaccination efforts are implemented in the host population.

**Figure 4:**
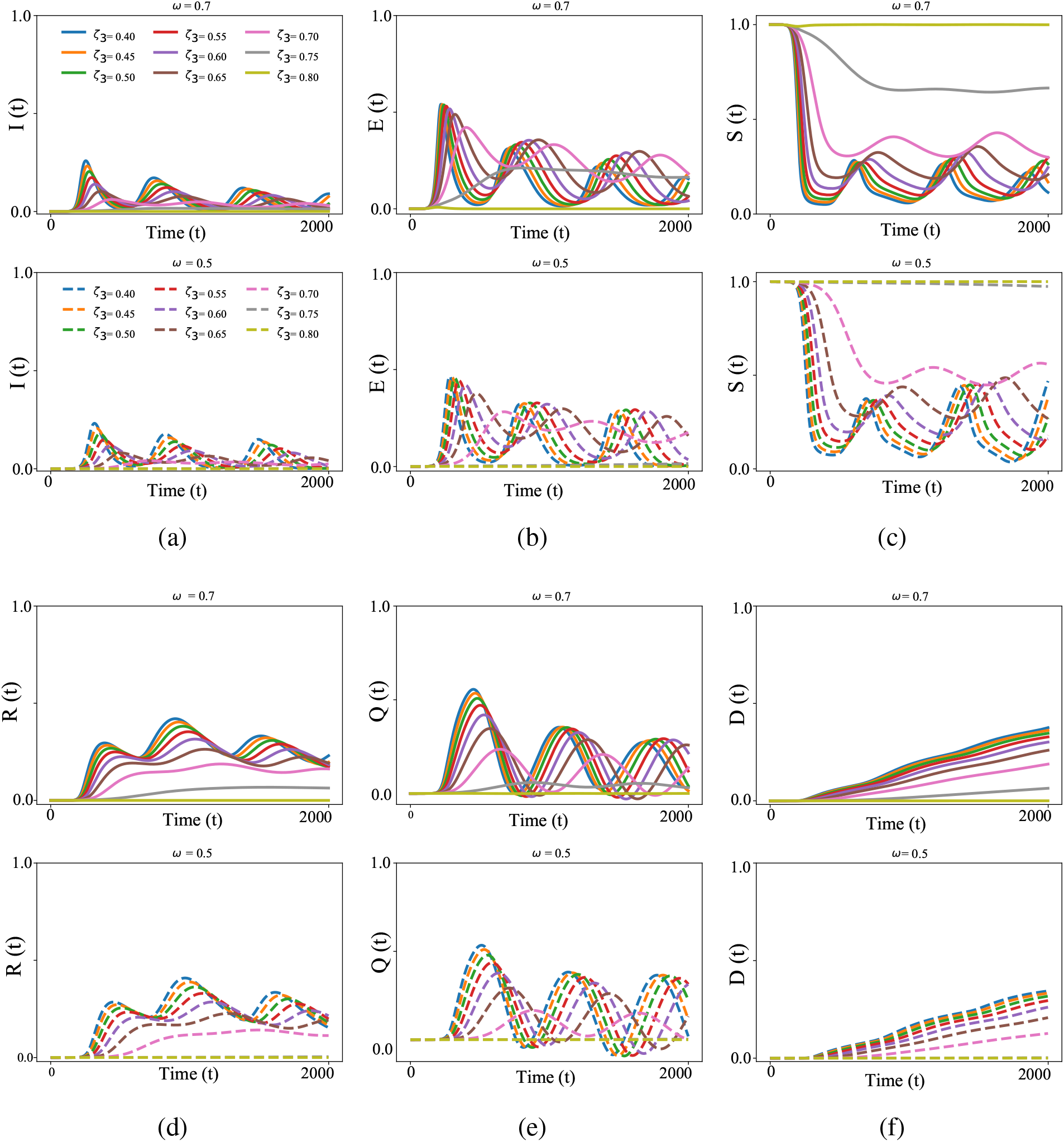
Simulation Results of the proposed temporal event-based probabilistic *SEIQRD* model (5), designed in this paper for various states when *ζ*_3_ ∈ [0.4, 0.5, 0.6, 0.7, 0.8] and *ω* ∈ [0.5, 0.7]. Figure.4a, 4c, 4b, 4d, 4e, 4f represents the evolution for infectious (*I*), susceptible (*S*), exposed (*E*), recovered (*R*), quarantined (*Q*) and dead states (*D*) in 2000 unit time span. Our simulation shows that the disease spreading is halted entirely when transmission rate, *p* → 0. Figure.3b, 3a and Figure.4a show that disease continues to circulate in the host population while *p*(*t*) *>* 0.

**Figure 5:**
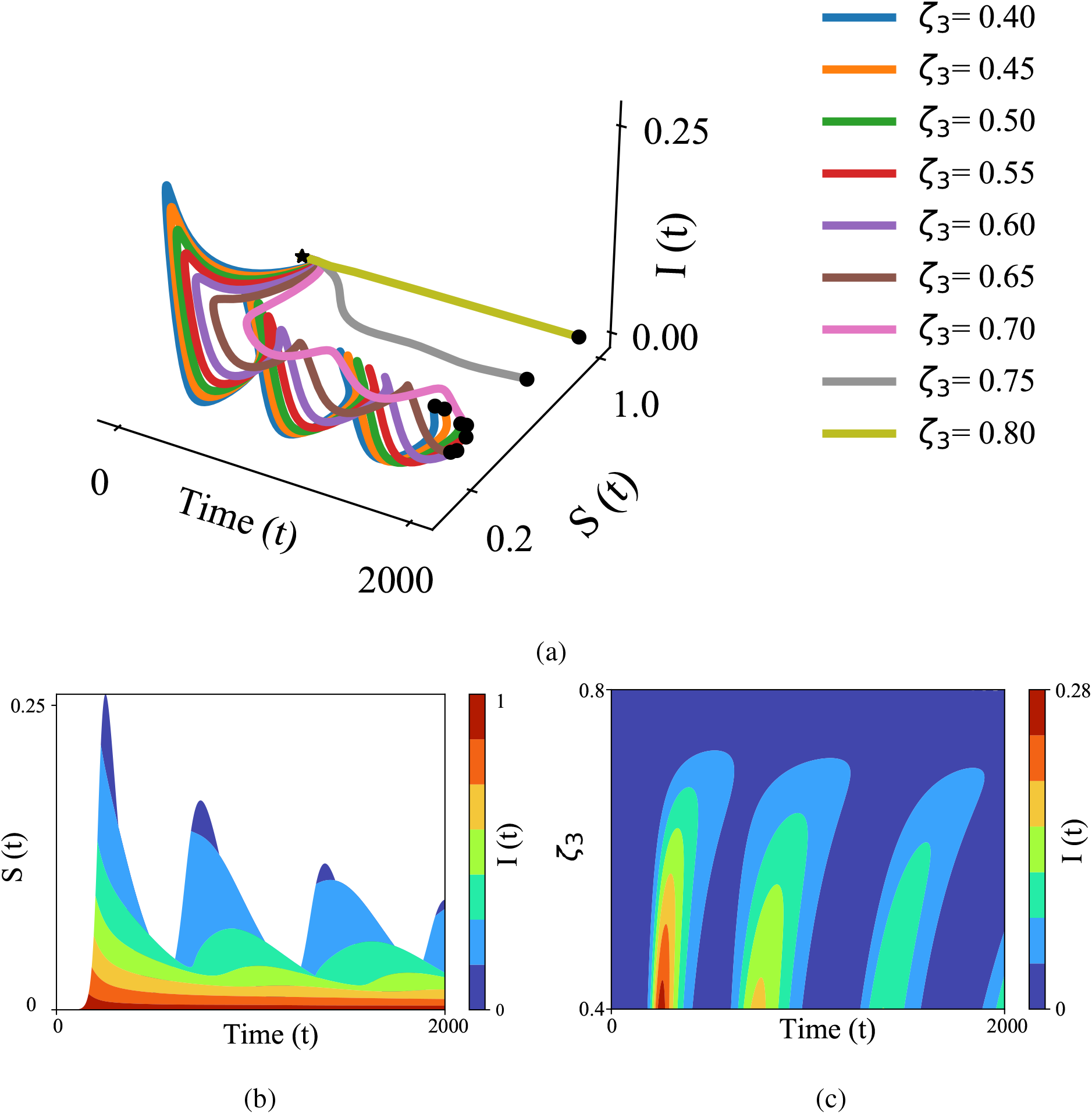
Trajectory of dynamical states *I* and *S* over time when *ω* = 7*e* − 1. Figure.5a: In this view, when *p*(*t*) *>* 0, the trajectories move outwards and diverge away which indicates an open epidemic state, meaning at any given time *t*, the communicable disease circulates in a subcluster of host population. In addition, a series of new cases and spikes are unavoidable due to waning immunity and recruitment to the susceptible population. Figure.5b: Size of infectious group (*I*) has an inverse relation with the size of susceptible group *S*. Figure.5c: Illustrates the relation between *ζ*_3_ and the size of infectious group (*I*). It shows that higher value of *ζ*_3_ results in fewer infectious cases. Therefore, the size of infectious group (*I*) has an inverse relation with *ζ*_3_.

**Figure 6:**
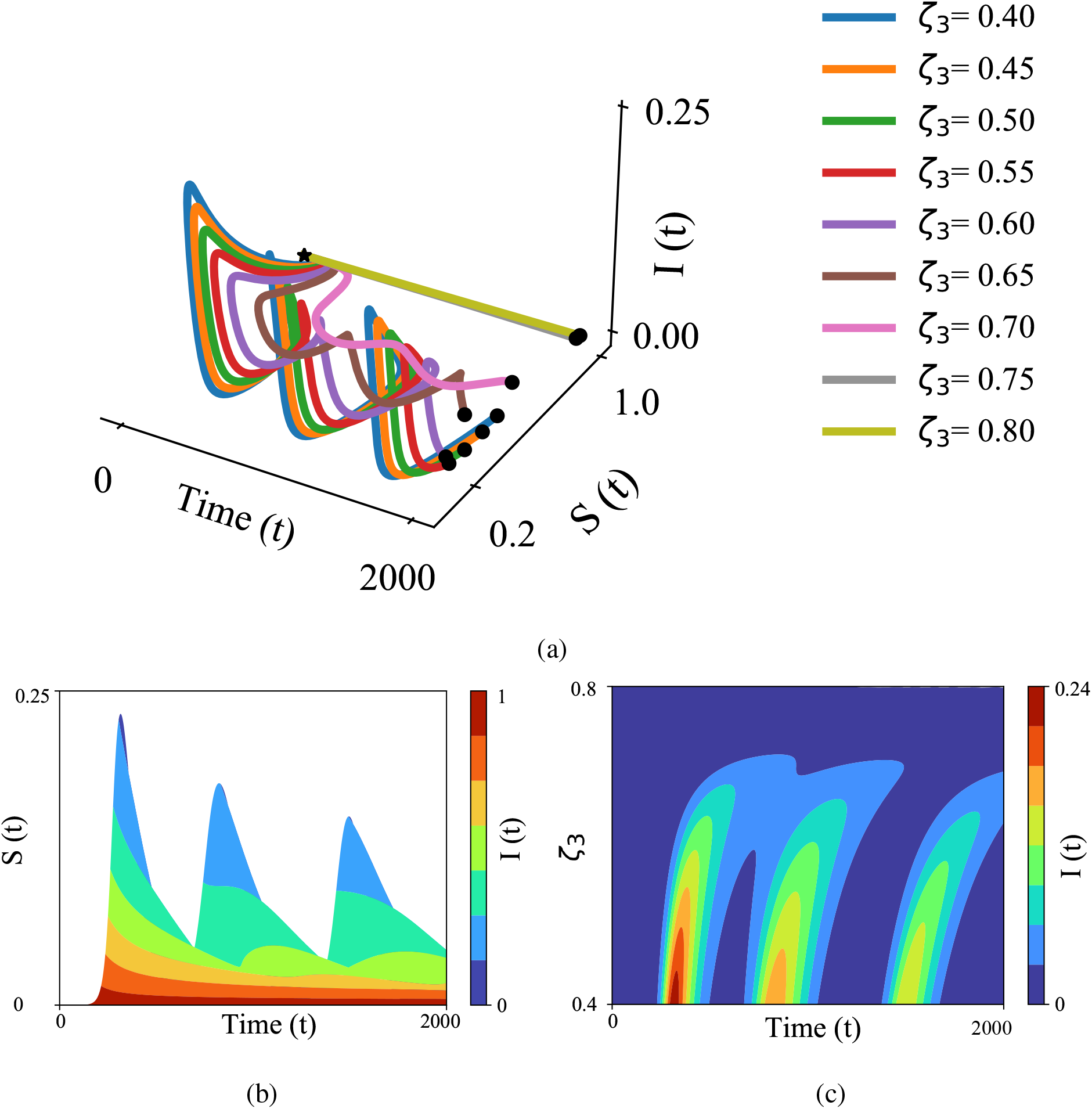
Trajectory of dynamical states *I* and *S* over time when *ω* = 0.5, using the proposed model of this paper. Reducing the contact rate, *ω*, in addition to increasing the immunity resilience, *v*(*t*), results in smaller infectious group size. Figure.6a: In this view, when *p*(*t*) *>* 0, the trajectories move outwards and diverge away which indicates an open epidemic state, meaning at any given time *t*,the communicable disease circulates in a sub-cluster of host population. In addition, a series of new cases and spikes are unavoidable due to waning immunity and recruitment to the susceptible population. Figure.6b: Size of infectious group (*I*) has inverse relation with size of susceptible group *S*. Figure.6c illustrates the relation between *ζ*_3_ and size of infectious group (*I*). It shows that higher value of *ζ*_3_ result in fewer infectious cases. Therefore, size of infectious group (*I*) has inverse relation with *ζ*_3_.

Figure.4 and 3 illustrate the evolution’s of model’s states and the interplay between 6 different states and *p*. We observe that number of recovered individuals (Figure.4d) non-monotonically increases and the fluctuations in the recovered population become more apparent for smaller value of *ζ*_3_ (which leads to larger value of *p*) due to insufficient immunity resilience against virus in the host population. Furthermore, insufficient immunity resilience against the virus has a similar effect on state *E* as indicated in Figure.4b. Particularly, insufficient immunity resilience and subsequently immunity loss both contribute to rate at which susceptible recruitment occur. Specifically, sufficient immunity resilience against virus leads to two scenarios: 1) outbreak fades away due to small number of infected individuals (*{ζ*_3_ = 8*e*^*−*1^, *ω* = 0.7*}* and *{ζ*_3_ *∈* [0.75, 0.8], *ω* = 0.5*}*) and 2) exposed individuals recruited to susceptible state at lower rate due to sufficient immunity resilience against virus (*{ζ*_3_ *∈* [0.7, 0.75], *ω* = 0.7*}* and *{ζ*_3_ *∈* [0.65, 0.7], *ω* = 0.5*}*).

To better understand the epidemic waves, we illustrated the trajectory of *I* as the function of time and *S* in Figure.5a and 6a. In this view, when *p*(*t*) *>>* 0, the trajectories move outwards and diverge away which indicates an open epidemic state, meaning at any given time *t*, the communicable disease circulates in a sub-cluster of host population. Furthermore, we observe that average contact rate, *ω*, impacts the rate at which system converges to endemic states. For example, when *ω* = 0.7, disease is halted entirely only when *ζ*_3_ = 0.8]. While for *ω* = 0.5 disease is eradicated when *{ζ*_3_ *∈* [0.75, 0.8]*}* (Figure.6 and 5). In addition, series of new cases and spikes are unavoidable due to waning immunity and recruitment to the susceptible population.

Figure.5c and 6c illustrate that reducing the contact rate (*ω* = 0.5) in addition to increasing the immunity resilience, *v*(*t*), results in smaller value of *p*(*t*) and eventually smaller infectious group size (*I*_*ω*=.5_ = 0.24 vs *I*_*ω*=.7_ = 0.28).

Our model framework allows robust prediction regarding disease spread in the presence of vaccination, waning immunity, and virus mutation. We find that halting the disease spread is achievable when the transmission rate converges to zero. Furthermore, our simulation shows that the rate at which *p* converges to zero is a product of host population’s average contact rate, immunity against virus, and virus mutation. Particularly, the rate at which immunity resilience is achieved in the host population compared to virus mutation occurrences defines the prevalence of spike in new cases. Our model significantly elucidates the interplay between immunity loss, virus mutation, immunity resilience, and prevalence of spike in new cases when exposed group is an intermediate step between the susceptible and the infectious groups. Furthermore, our model maps out the endemic state given contributing factors and predicts the size of epidemic waves.

## Materials

In this paper, we propose a new computational model using a homogeneous Poisson point process for the first time which addresses the critical issues pertaining to conventional state-space infectious disease modeling. The proposed model in this paper reconstructs the *S → E → I* transition, by considering the fact that changes in the susceptible population occur only when individuals get exposed to the virus and not when they contract the disease. Furthermore, our proposed framework takes into account the probabilistic nature of transition and the temporal inter-dependency between *p, ω, S*, and *I* when an exposed individual undergoes an incubation period. In addition, our proposed model has the flexibility to take into account mutation, development of immunity resilience, and immunity loss as part of the state-space model.

This unique framework allows for the evaluation of the disease spread in various societies and sub-societies with different immunity responses and vaccination profiles. Our proposed model provides a robust framework that can be imperative when controlling a long pandemic in a mega population that echoes waves of mutation and spread. Furthermore, Such a model can be utilized to simulate the future waves of pandemics depending on an assumed temporal expectation of the mutation.

Our model provides a unique mathematical framework that allows policy-makers to improve preparedness and unfolds the optimal time-frame for vaccination given the available resources for the current and unforeseen outbreaks. Using our framework, health authorities will have a powerful and flexible framework to objectively conceptualize and predict the endemic state in the context of mitigation interventions and deployment of vaccination.

### Homogeneous Poisson Point Processes

A homogeneous Poisson point process is a stochastic process that is utilized in queuing theory to model random events such as arrivals or departures in a system (47). The Poisson point process is defined as a Poisson random variable where the Poisson parameter depends on the duration of the interval in which departure or arrival occurs. In Poisson point process, non-overlapping intervals are considered as independent events (48) (49). Considering these two key observations, a Poisson point process is defined as given below.

#### Definition 0.1.

Assume *X*(*t*) = *Z*(*t*_1_, *t*_2_) represents a Poisson point process. The number of arrivals, *k*, during (*t*_1_, *t*_2_) interval with length of *t* = *t*_2_–*t*_1_ is a Poisson random variable with parameter *λt*. Given that,

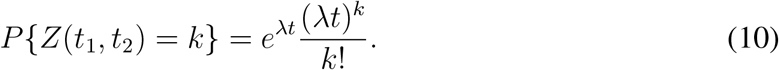

where *P {Z*(*t*_1_, *t*_2_) = *k}* represents probability of having *k* arrival within *t* unit time. Considering (10), it can be mentioned that if the intervals (*t*_1_, *t*_2_) and (*t*_3_, *t*_4_) are non-overlapping, then the random variables *Z*(*t*_1_, *t*_2_) and *Z*(*t*_3_, *t*_4_) are independent.•

The properties of a Poisson process imply that in any interval *δ*(*t*), one event can occur with the probability that is proportional to *δ*(*t*). Furthermore, the probability that two or more events occur in the same interval is proportional to *O*(*δ*(*t*)) (50). The inter-arrival duration of a Poisson point process (inter-arrival duration between the (*i −* 1)^*th*^ and (*i*)^*th*^ moments) is defined as an exponential process. The aforementioned statement is proven below.

*Proof*. Assume *t*_0_ is any fixed point and *t*_0_+*τ* represents the first arrival time after *t*_0_. Therefore, the probability of having at least one arrival within *τ* unit time, *F*_*τ*_ (*t*), is:

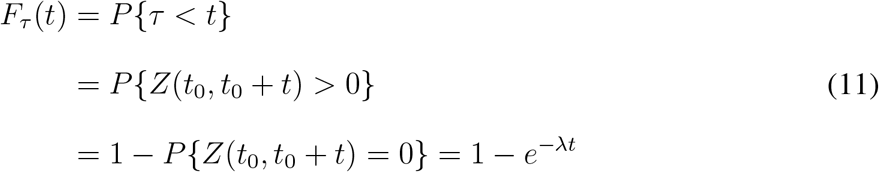

We can observe that 1 *− e*^*−λt*^ is in fact, the cumulative distribution function of exponential distribution. Hence, we can derive the probability density function (*PDF*) as follows:

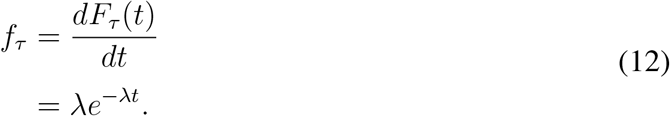

Thus, considering (12), we can derive the average number of arrivals in a Poisson point process given a known interval time *τ* as follows (49) (51):

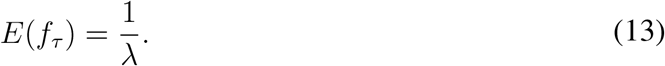

We use the Poisson point process concept presented in this section to reformulate the conventional definition of the *SEIR* model. In the next section, we propose a novel *SEIR* framework by integrating the arrival Poisson point process concept into the *SEIR* state-space model. Thus introducing a coherent framework that takes into account the interdependency between *ω, p*, and *I* state that are neglected in the conventional *SEIR* model.

## Data Availability

All data produced in the present study are available upon reasonable request to the authors

## Acknowledgement

The material presented in this work is supported by US National Science Foundation (NSF), Awards 2208189, and 2031594.

## References

1. H. Association, et al., Hospitals and health systems continue to face unprecedented financial challenges due to covid-19, Published June (2020).

2. G. Giordano, et al., Modelling the covid-19 epidemic and implementation of population-wide interventions in italy, Nature medicine 26, 855–860 (2020).

3. L. Luo, et al., Modes of contact and risk of transmission in covid-19: a prospective cohort study 4950 close contact persons in guangzhou of china (2020).

4. S. Asadi, N. Bouvier, A. S. Wexler, W. D. Ristenpart, The coronavirus pandemic and aerosols: Does covid-19 transmit via expiratory particles? (2020).

5. T. K. Chan, Universal masking for covid-19: evidence, ethics and recommendations, BMJ Global Health 5, e002819 (2020).

6. J. Chen, R. Wang, N. B. Gilby, G.-W. Wei, Omicron (b. 1.1. 529): Infectivity, vaccine breakthrough, and antibody resistance, ArXiv (2021).

7. N. L. Miller, T. Clark, R. Raman, R. Sasisekharan, Insights on the mutational landscape of the sars-cov-2 omicron variant, bioRxiv (2021).

8. S. Collie, J. Champion, H. Moultrie, L.-G. Bekker, G. Gray, Effectiveness of bnt162b2 vaccine against omicron variant in south africa, New England Journal of Medicine (2021).

9. W. F. Garcia-Beltran, et al., mrna-based covid-19 vaccine boosters induce neutralizing immunity against sars-cov-2 omicron variant, Cell (2021).

10. L.-S. Young, S. Ruschel, S. Yanchuk, T. Pereira, Consequences of delays and imperfect implementation of isolation in epidemic control, Scientific reports 9, 1–9 (2019).

11. I. Siettos, L. Russo, Mathematical modeling of infectious disease dynamics, Virulence 4, 295–306 (2013).

12. J. A. McCullers, J. D. Dunn, Advances in vaccine technology and their impact on managed care, Pharmacy and Therapeutics 33, 35 (2008).

13. R. Rappuoli, M. Pizza, G. Del Giudice, E. De Gregorio, Vaccines, new opportunities for a new society, Proceedings of the National Academy of Sciences 111, 12288–12293 (2014).

14. E. P. Hoberg, D. R. Brooks, Evolution in action: climate change, biodiversity dynamics and emerging infectious disease, Philosophical Transactions of the Royal Society B: Biological Sciences 370, 20130553 (2015).

15. J. R. Rohr, et al., Emerging human infectious diseases and the links to global food production, Nature sustainability 2, 445–456 (2019).

16. R. Brooks, E. P. Hoberg, W. A. Boeger, The Stockholm paradigm: climate change and emerging disease (University of Chicago Press, 2019).

17. A. James, et al., Successful contact tracing systems for covid-19 rely on effective quarantine and isolation, medRxiv (2020).

18. D. Lewis, Why many countries failed at covid contact-tracing-but some got it right., Nature pp. 384–387 (2020).

19. M. J. Ryan, T. Giles-Vernick, J. E. Graham, Technologies of trust in epidemic response: openness, reflexivity and accountability during the 2014–2016 ebola outbreak in west africa, BMJ Global Health 4, e001272 (2019).

20. W. E. R. Team, Ebola virus disease in west africa—the first 9 months of the epidemic and forward projections, New England Journal of Medicine 371, 1481–1495 (2014).

21. B. Ivorra, M. R. Ferrández, M. Vela-Pérez, A. M. Ramos, Mathematical modeling of the spread of the coronavirus disease 2019 (covid-19) taking into account the undetected infections. the case of china, Communications in nonlinear science and numerical simulation 88, 105303 (2020).

22. J. Ma, Z. Ma, Epidemic threshold conditions for seasonally forced seir models, Mathematical Biosciences & Engineering 3, 161 (2006).

23. Y. N. Kyrychko, K. B. Blyuss, Global properties of a delayed sir model with temporary immunity and nonlinear incidence rate, Nonlinear analysis: real world applications 6, 495–507 (2005).

24. R. Li, C. J. E. Metcalf, N. C. Stenseth, O. N. Bjørnstad, A general model for the demographic signatures of the transition from pandemic emergence to endemicity, Science Advances 7, eabf9040 (2021).

25. A. Radulescu, C. Williams, K. Cavanagh, Management strategies in a seir-type model of covid 19 community spread, Scientific reports 10, 1–16 (2020).

26. O. N. Bjørnstad, K. Shea, M. Krzywinski, N. Altman, The seirs model for infectious disease dynamics., Nature Methods 17, 557–559 (2020).

27. N. Hens, et al., Seventy-five years of estimating the force of infection from current status data, Epidemiology & Infection 138, 802–812 (2010).

28. C. C. McCluskey, Global stability for an seir epidemiological model with varying infectivity and infinite delay, Mathematical Biosciences & Engineering 6, 603 (2009).

29. G. Huang, Y. Takeuchi, W. Ma, D. Wei, Global stability for delay sir and seir epidemic models with nonlinear incidence rate, Bulletin of mathematical biology 72, 1192–1207 (2010).

30. L. Gallo, M. Frasca, V. Latora, G. Russo, Lack of practical identifiability may hamper reliable predictions in covid-19 epidemic models, Science advances 8, eabg5234 (2012).

31. S. Unkel, C. P. Farrington, P. H. Garthwaite, C. Robertson, N. Andrews, Statistical methods for the prospective detection of infectious disease outbreaks: a review, Journal of the Royal Statistical Society: Series A (Statistics in Society) 175, 49–82 (2012).

32. R. E. Serfling, Methods for current statistical analysis of excess pneumonia-influenza deaths, Public health reports 78, 494 (1963).

33. C. Farrington, N. J. Andrews, A. Beale, M. Catchpole, A statistical algorithm for the early detection of outbreaks of infectious disease, Journal of the Royal Statistical Society: Series A (Statistics in Society) 159, 547–563 (1996).

34. S. C. Wieland, J. S. Brownstein, B. Berger, K. D. Mandl, Automated real time constant-specificity surveillance for disease outbreaks, BMC medical informatics and decision making 7, 1–10 (2007).

35. J. Zhang, F.-C. Tsui, M. M. Wagner, W. R. Hogan, AMIA Annual Symposium Proceedings (American Medical Informatics Association, 2003), vol. 2003, p. 748.

36. Y. Le Strat, F. Carrat, Monitoring epidemiologic surveillance data using hidden markov models, Statistics in medicine 18, 3463–3478 (1999).

37. Madigan, Bayesian data mining for health surveillance, Spatial and syndromic surveillance for public health pp. 203–221 (2005).

38. S. Dahal, J. M. Banda, A. I. Bento, K. Mizumoto, G. Chowell, Characterizing all-cause excess mortality patterns during covid-19 pandemic in mexico, BMC Infectious Diseases 21, 1–10 (2021).

39. P. B. Miller, E. B. O’Dea, P. Rohani, J. M. Drake, Forecasting infectious disease emergence subject to seasonal forcing, Theoretical Biology and Medical Modelling 14, 1–14 (2017).

40. Z. Li, et al., Impact of a two-dose varicella immunization program on the incidence of varicella: a multi-year observational study in shanghai, china, Expert Review of Vaccines 20, 1177–1183 (2021).

41. R. Paul, A. A. Arif, O. Adeyemi, S. Ghosh, D. Han, Progression of covid-19 from urban to rural areas in the united states: a spatiotemporal analysis of prevalence rates, The Journal of Rural Health 36, 591–601 (2020).

42. W. O. Kermack, A. G. McKendrick, A contribution to the mathematical theory of epidemics, Proceedings of the royal society of london. Series A, Containing papers of a mathematical and physical character 115, 700–721 (1927).

43. J. Satsuma, R. Willox, A. Ramani, B. Grammaticos, A. Carstea, Extending the sir epidemic model, Physica A: Statistical Mechanics and its Applications 336, 369–375 (2004).

44. M. J. Keeling, P. Rohani, Modeling infectious diseases in humans and animals (Princeton university press, 2011).

45. L. Dell’Anna, Solvable delay model for epidemic spreading: the case of covid-19 in italy, Scientific Reports 10, 1–10 (2020).

46. S. Tiwari, C. Vyasarayani, A. Chatterjee, Data suggest covid-19 affected numbers greatly exceeded detected numbers, in four european countries, as per a delayed seiqr model, Scientific reports 11, 1–12 (2021).

47. L. Kleinrock, Queueing systems: theory (John Wiley, 1975).

48. R. G. Gallager, Discrete stochastic processes, vol. 321 (Springer Science & Business Media, 2012).

49. A. Papoulis, S. Pillai, Probability, random variables, and stochastic process, mcgraw-hill, inc (1991).

50. J. H. Jones, Notes on r0, California: Department of Anthropological Sciences 323, 1–19 (2007).

51. J. Pitman, Poisson-kingman partitions, Lecture Notes-Monograph Series pp. 1–34 (2003).

